# Acute cardiac injury in patients with COVID-19

**DOI:** 10.1101/2020.05.18.20105866

**Authors:** Andrea De Lorenzo, Daniel Arthur B. Kasal, Bernardo Rangel Tura, Cristiane da Cruz Lamas, Helena Cramer Veiga Rey

**Author notes:** Corresponding author, Instituto Nacional de Cardiologia, Research and Teaching Department, Rua das Laranjeiras, 374, 22240-006, Rio de Janeiro, Brazil.

## Abstract

**Introduction:** Cardiac complications of COVID-19 are potentially life-threatening. The occurrence of myocardial injury in the context of COVID-19 is multifactorial and has generated increasing interest.

**Methods:** A systematic review with meta-analysis of the literature was performed. MEDLINE and EMBASE were searched. Two independente reviewers evaluated the selected manuscripts for the outcome “myocardial injury”, defined by troponin elevation above the 99th percentile. Study heterogeneity and risk of bias were evaluated.

**Results:** Eight studies, with a total of 1229 patients, were included. The frequency of myocardial injury was 16% (95% CI: 9% - 27%). The heterogeneity among studies was high (93%). Conclusions: Myocardial injury may occur in patients with COVID-19, with a frequency of 16% among current studies. Continuous research is needed to update these findings, as the pandemic evolves, and to define the implications of myocardial injury in the context of this infection.

## Introduction

The association between cardiovascular disease (CVD) and adverse outcomes in patients with COVID-19 has been consistently described. In the series of Ruan *et al*, among 68 patients who died, 19% had CVD, versus none among those who survived (p<0,001) [1]. However, there is also evidence of acute cardiac damage in the context of the infection. Recently, the first proven case of direct cardiac damage was published, with demonstration of viral particles inside cardiomyocytes of a patient with COVID-19 with cardiogenic shock who underwent endomyocardial biopsy [2].

Troponin elevations, suggesting myocardial damage, are frequent in severe infections [3]. Abnormal troponin values have been frequently found in patients with COVID-19, especially when high-sensitivity troponin is employed. Among the first 41 COVID-19 patients in China, 5 (12%) had acute myocardial injury (manifested by troponin elevations > 99th percentile), and 4 of these needed intensive care treatment [4], demonstrating the increased severity associated with this presentation. The aim of this study was to therefore evaluate the frequency of acute myocardial injury in patients with COVID-19.

## Methods

A systematic review of the literature was performed to evaluate the frequency of acute myocardial injury expressed by abnormal troponin levels (>99th percentile) in hospitalized patients with COVID-19.

### Population

Inpatients with confirmed COVID-19.

### Exposure

Exposure to SARS-CoV-2, with clinical COVID-19.

### Outcomes

Abnormal troponin levels (>99th percentile), with or without clinical manifestation of acute cardiac injury.

### Study types

Case series or cohorts.

### Sources of information

Two databases were searched: MEDLINE and Embase. Other manuscripts were manually added from reviews or other sources of data.

### Search strategies

Search strategies are described in the Supplementary Material. The search was performed until 04/02/2020. No language restrictions were applied.

### Selection strategy

Manuscripts were selected according to title, abstract and full text and were analyzed by two independent reviewers. Discordances were solved by a third reviewer. The quality of the studies was graded according to the Newcastle-Ottawa scale.

### Statistical analysis

A meta-analysis of random effects was performed by the DerSimonian and Laird method, and results were shown in forest plots. The measure of effect was the logit of the proportion of myocardial injury. Heterogeneity was estimated by I^2^. In the presence of substantial (>50%) heterogeneity, a meta-regression was performed using clinical and demographic variables as determinants. The presence of bias was analyzed using a funnel plot and also quantitatively by Egger’s test.

## Results

The PRISMA flowchart is shown in the Supplementary Material. After exclusions, eight manuscripts were considered eligible for the meta-analysis. The overall quality of the manuscripts was graded as low to moderate.

### Acute myocardial injury

Table 1 depicts the studies which described the occurrence of cardiac injury in hospitalized COVID-19 patients. The frequencies of myocardial injury ranged from 3.3% to 44.4% [1,4-10]. Figure 2 shows the results of the meta-analysis. In hospitalized patients with COVID-19, the frequency of myocardial injury was 16% (95%CI: 9%-27%). There was high heterogeneity among the studies (93%), so a meta-regression was performed (Table 2). Age and the presence of chronic obstructive pulmonary disease (COPD) were found as a sources of heterogeneity, and then another meta-analysis was performed with stratification for these variables (Figure 3). The studies with outlier values (Ruan *et al* [1] and He *et al* [5]) were analyzed separately, resulting in different frequencies of myocardial injury, with 33% in the worst scenario; however, heterogeneity was still high (92% before stratification and 63% after, respectively).

**Table 1.** Frequency of acute myocardial injury in patients with COVID-19

| Study | N | Acute myocardial injury |
| --- | --- | --- |
| Huang *et al*<sup>4</sup> | 41 | 12.0% |
| He *et al*<sup>5</sup> | 54 | 44.4% |
| Wang *et al*<sup>6</sup> | 138 | 7.2% |
| Shi *et al*<sup>7</sup> | 416 | 19.7% |
| Ruan *et al*<sup>1</sup> | 150 | 3.3% |
| Guo *et al*<sup>8</sup> | 187 | 27.8% |
| Zhou *et al*<sup>9</sup> | 191 | 17.0% |
| Yang *et al*<sup>10</sup> | 52 | 23.0% |

**Figure 2.**
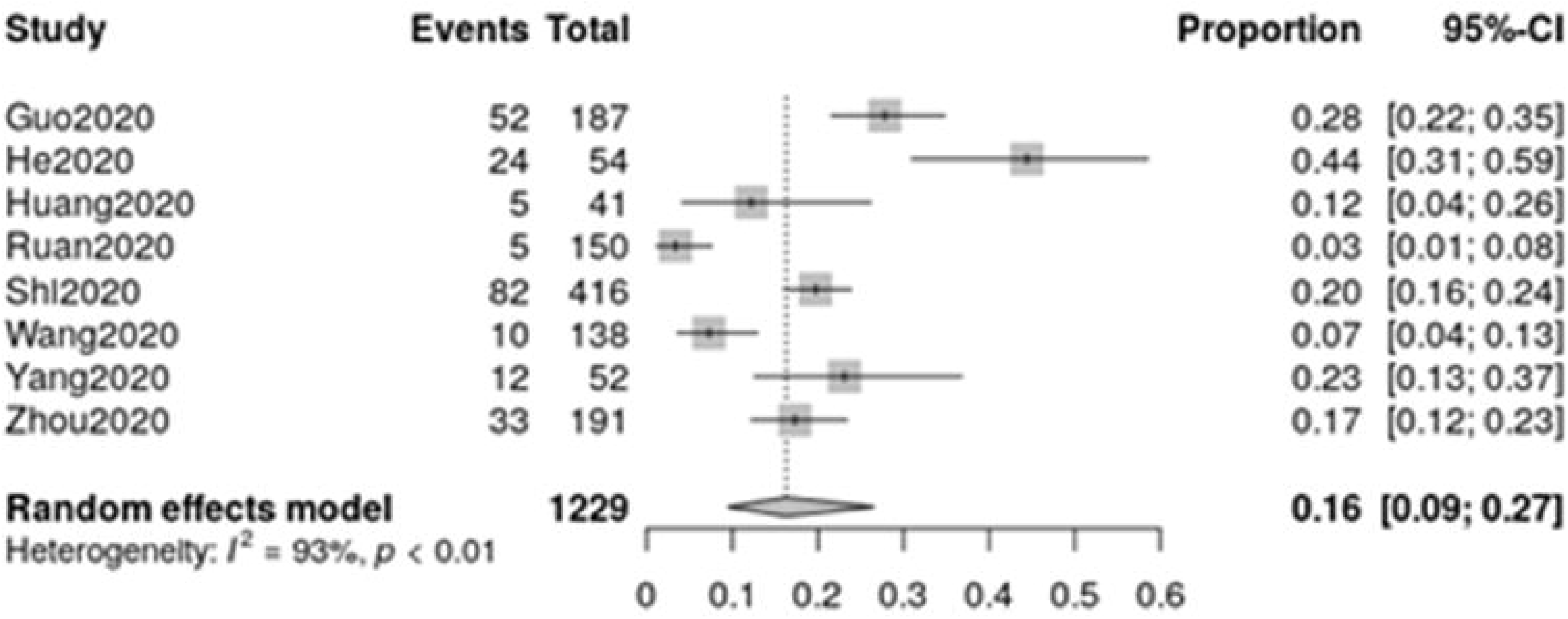
Meta-analysis of the frequency of myocardial injury

**Table 2.** Meta-regression for the analysis os heterogeneity among studies

|  | Coefficient | Explained heterogeneity | P value |
| --- | --- | --- | --- |
| Age | 0,0889 | 2.35% | 0.029 |
| Male gender | -0.0007 | 0 | 0.98 |
| Hypertension | 0.0496 | 20.25% | 0.138 |
| Diabetes | -0.0556 | 5.43% | 0.238 |
| Prior cardiovascular disease | 0.0498 | 1.02% | .,068 |
| Chronic obstructive pulmonary disease | 0.1048 | 30.66% | 0.016 |

**Figure 2.**
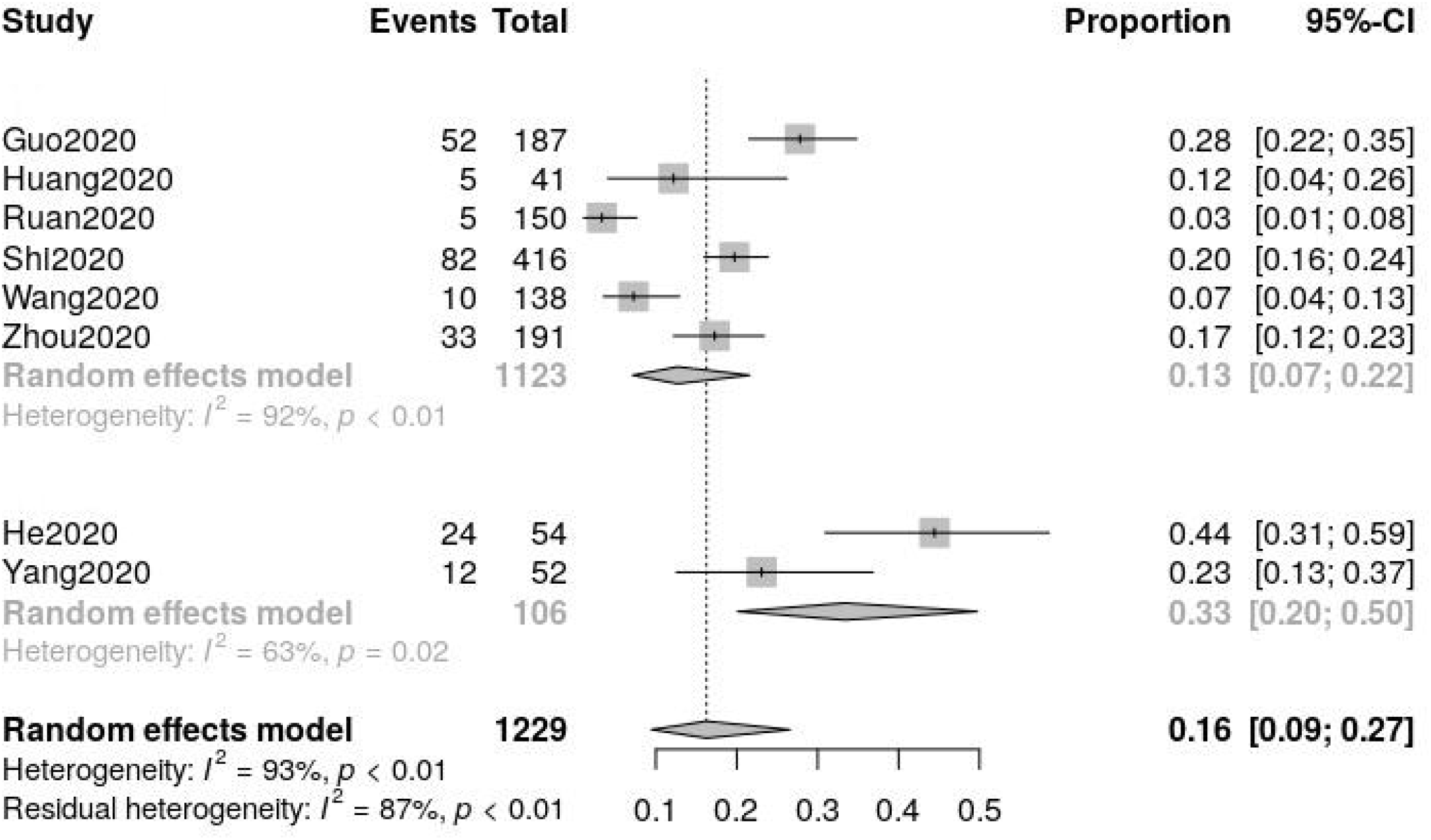
Stratified meta-analysis for the evaluation of heterogeneity

**Figure 3.**
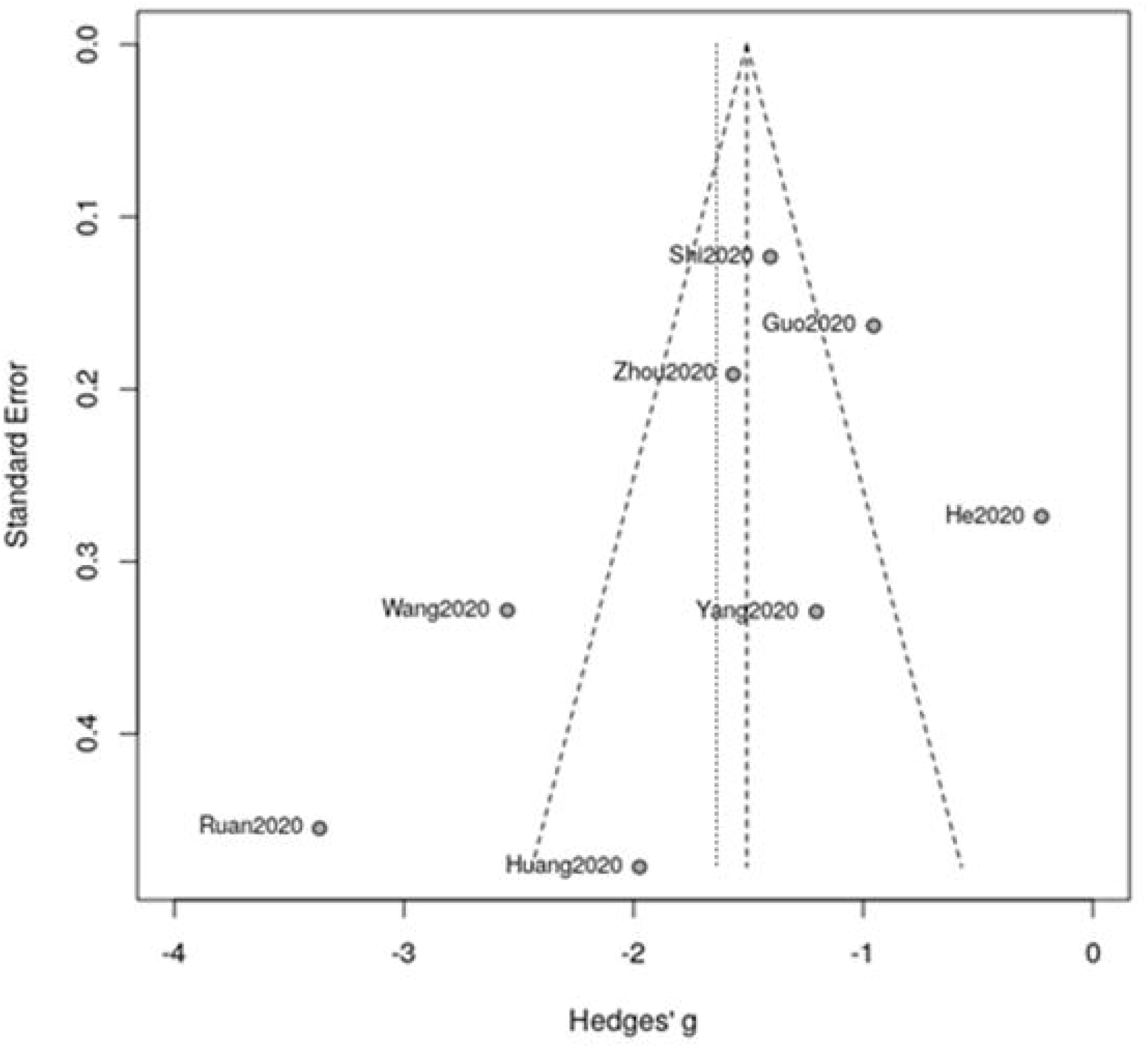
Funnel plot for the evaluation of publication bias

Regarding publication bias (Figure 4), even though some asymmetry was observed on the funnel plot, there was no significant bias (p = 0.3418).

## Discussion

In the face of the COVID-19 pandemic – an unprecedented situation in modern Medicine – it is of key importance to learn about the characteristics of the disease and its complications. Cardiac injury in COVID-19 is a main complication which should be accounted for in patients who clinically deteriorate, and anticipated in those with risk factors such as prior cardiovascular disease. Adequate comprehension of its frequency, predisposing factors and characteristics is therefore desirable.

The definition of acute cardiac injury is controversial, as most studies consider the elevation of troponin above the 99th percentile as the criterion for myocardial damage, without necessarily any association with clinical, electrocardiographic or imaging data. Additionally, different sensitivities of tests and cutpoints for abnormalities may hamper the interpretation of the studies. Of note, the timing of the troponin measurements was different among the studies, and patient severity was also variable. These may explain the different frequencies of myocardial injury between the studies; nonetheless, the overall frequency of 16% depicts an important issue, which should be underscored.

The mechanisms of myocardial injury in COVID-19 may include atherosclerotic plaque rupture and tipe I acute myocardial infarction, supply/demand imbalance in the context of severe infection and sepsis, with type II myocardial infarction, or direct myocardial damage, as demonstrated by Tavazzi *et al* [2], due to the pathophysiologic substrate (ACE-II receptor) in cardiomyocytes [11]. All these may be precipitated or aggravated by systemic inflammation and the “cytokine storm” [1]. Abnormal troponin values, especially if high-sensitivity troponin is employed, have been frequently reported in patients with COVID-19 since the beggining of the pandemic. Among the first 41 Chinese patients with COVID-19, 5 (12%) had acute myocardial injury, and 4/5 needed intensive care support. [4]. Clinical data are key to distinguish the different disease entities, as shown in case reports; for example, Hu et al [12] described the case of a 37-year old male, admitted with chest pain and shortness of breath, who was diagnosed with COVID-19 and displayed ST-segment elevation in inferior leads, troponin T > 10.000 ng/L, BNP of 21.025 ng/L, but had no coronary stenoses on the CT angiogram. The echocardiogram revealed severe left ventricular dysfunction, and the final diagnosis was of fulminant myocarditis with cardiogenic shock. Therefore, it should be emphasized that abnormal troponin results may be multifactorial and should be interpreted carefully and in conjunction with other data, due to thir low specificity for the diagnosis of acute myocardial infarction. Nonetheless, as Chapman *et al* [13] suggest, troponin may also be viewed as an ally, if emphasis is placed on its prognostic value (and not on the diagnosis of acute coronary artery disease), and also regarding the epidemiologic issue of recognizing the overall incidence of myocardial injury in COVID-19 patients. Even so, it must be underscored that abnormal troponin levels should not be considered an indication for procedures such as cardiac catheterization, nor for therapeutic measures such as the initiation of antiplatelet agentes.

Finally, the current speed of release of data and new publications makes it especially hard to review the literature, which is fastly evolving. However, a meta-analysis may be useful to provide an estimate of the frequency of events, which in the present case constitute a particularly dangerous complication, which should be kept in mind.

## Conclusions

In the context of a severe infection, the implications of abnormal troponin measurements are various, and elevated troponin in COVID-19 may not imply the occurrence of acute myocardial infarction or myocarditis. Nonetheless, the present review shows that myocardial injury may occur in 16% of the hospitalized patients with COVID-19, a frequency which is high enough to draw attention to this complication.

## Data Availability

All data referred to in the manuscript are available (the manuscript is a meta-analysis of current literature).

## Acknowledgements

The authors thank Braulio Santos for discussing the results, Francijane Oliveira and Cyntia Aguiar for data search, and Marcelo Goulart for the support with the elaboration of figures.

